# Influence of Malaria Edemicity and TB Prevalence / BCG Coverage on COVID-19 Mortality

**DOI:** 10.1101/2020.09.09.20191684

**Authors:** Tareef Fadhil Raham

**Author notes:** 009647901584338.

## Abstract

**Background:** Regarding SARS-CoV-2 it is well known that a substantial percentage of adult population cannot get infected if exposed to this novel coronavirus. Several studies give primary indication about the possible role of preexisting immunity whether cross immunity or not. Possible role of latent TB, BCG and malaria have been already suggested to create innate cross heterogeneous immunity. We look for influence of these factors on Covid-19 mortality in malarious countries.

**Material and methods:** 80 malarious countries were enrolled in this study. Hierarchical multiple regression type of analysis was used for data analyses. TB prevalence/ 100,000 population standardized to BCG coverage rates was taken as direct factor in the test. Malaria incidence /1000 population was considered as intermediate factor and the outcome was COVID-19 mortality/ 1 million (M) population.

**Results:** Hierarchical multiple regression analysis showed significant associations between standardized TB prevalence to BCG coverage and reduced COVID-19 mortality. This analysis also showed that malaria have an additional effect in reducing COVID-19 mortality with high significant association too.

**Conclusions:** Malaria and standardized TB prevalence are statistical significant factors predicting COVID-19 mortality in negative associations.

## Introduction

Microorganisms infecting mammalian hosts may modulate long lasting protective heterologous cross-immunological reactions once exposed to heterologous agonists in the future ^1,2,3^. In 2011, Netea *et al* proposed the term “trained immunity” to describe this ability of innate immune cells to nonspecifically adapting, protecting and remembering primary stimulation^4^.

### BCG

BCG through attenuated strain of *Mycobacterium bovis* was used to produce heterogeneous immunity against *Mycobacterium tuberculosis* which is another species within the genus *Mycobacterium*. BCG was blamed to lead to incomplete and often varying degree of protection against TB disease^5^. On the other hand, studies have been done over the past few decades showed that certain adaptations connected with innate immune cells (monocyte/macrophages, NK (Natural Killer) cells are responsible for nonspecific effects of vaccination beyond its target ^6,7^.These studies showed that Vaccination with BCG induces an improved innate immune response against microorganisms other than Mycobacterium, this includes bacteria such as *Staphylococcus aureus*, fungi such as *Candida albicans* and viruses such as the yellow fever virus ^6,8,9^.

There is well known evidence that BCG plays a role in reduction of neonatal sepsis and respiratory infection reduction^10,11,12^. Its role in childhood mortality reduction is known since 1927^13,14,15^. Furthermore BCG vaccination was associated with diminished morbidity and mortality rates associated with malaria, unclassified fever, preventing, sepsis and leprosy ^10,11,13,16,17,18^. In spite of that many countries discontinue BCG vaccinations or limited its use to high risk groups because great achievements in reduction of TB prevalence rates.

### TB

Tuberculosis (TB) is one of the major cause of illness and death in many countries and a significant public health problem worldwide. TB disease is one of the top 10 causes of death and accounted to estimated 10 million people in 2018^19^.

Latent tuberculosis infection (LTBI) is the persistence of an immunological response to *Mycobacterium tuberculosis* antigen stimulation without any clinically active disease^20^. The WHO recommends tailored latent tuberculosis infection management based on tuberculosis burden and resource availability^21^. Treatment of active disease is by far the major intervention in this regard worldwide, while diagnosis and treatment of LTBI are hindered by the cost implications of testing, lack of a consensus on the tests recommended, and side effects of treatment^22^.

Treatment of LTBI in low prevalence (high to upper-middle-income) countries like USA^23^and many European countries^24^is feasible (in spite of prolonged adaptive immunological response to *Mycobacterium* antigen), as elimination of this reservoir of infection will reduce the burden of the disease. However, the scenario in high prevalence countries like many countries in Asia and Africa is quite the opposite, where reinfection due to contact with active cases rather than reactivation contributes to a high disease burden^24^. Latent TB infections was suggested to create heterogeneous immune response to COX-COV2 viral infection in different patterns and severity according to different BCG status ^25^ further studies consolidate such latent TB role thereafter ^26,27,28^.

### Malaria

Malaria infection can be asymptomatic, or, more accurately, “subclinical,” because subtle symptoms and chronic health effects may occur but not lead to clinical diagnosis and treatment^29^. In similar way for TB control, malaria elimination requires eradicating both clinically symptomatic as well as these “silent” infections^30^ which may serve as a reservoir and contribute to ongoing malaria transmission^31,32^.

Malarial infection cross protection for other microorganisms is well known for *Mycobacterium tuberculosis*. The prevalence of malaria/TB coinfection is low^33^. Murine models suggested protective humoral and cellular immune responses against the complications of each disease]^34,35.^ Further studies suggested increased production of gamma interferon (IFN-γ), tumor necrosis factor alpha (TNF-α), and humoral factors induced by tuberculosis infection that are protective against malaria infection^36,37, 36^.

Malaria was also founded to clear S. pneumonia much more efficiently in co-infected model ^37^. Furthermore, having malaria infection was associated with a reduced likelihood of influenza virus and vice versa^38,39^. It was also suggested that measles infection repressed parasitemia in addition to influenza A infection^40^. Heterogeneous immunological response also exists among different malaria species.

In Italy it has been thought that blacks are less affected by COVID-19 and due to suggested previous exposure to malaria and presence of anti-glycosylphosphatidylinositol antibodies (which have possible protective effect against malaria re-infection^41^) might give cross protection against COX-COV2 infection^42^. Furthermore epidemiological paradox between coronavirus disease 2019 (COVID-19) and malaria endemicity was noticed in the initial phase of this pandemic^43^.

Added to these evidence further study showed that high endemicity of TB and malaria, and universal BCG programs was suggested to have a cushioning effect on proportion of affected population by COVID-19 30.

Substantial percentage of adult population cannot get infected if exposed to SARS-CoV-2 is known that a ^44^. Several studies suggest a possible role of preexisting immunity whether cross immunity or not (antibodies/T cells, etc.) as a factor explaining such diversity^46^.

Both of BCG implementation and latent TB prevalence later on did not fully explain variances in COVID-19 mortalities across different countries. In South Africa for example, where COVID-19 mortality/M population is 238 / M at the study time, while TB prevalence/1000 population is 520/ 100,00 population which is very high. Another examples, these studies did not explain low COVID-19 mortality in countries which have relatively low TB prevalence/ 100,000 population as in : Togo, Benin and Mali where estimated TB prevalence of 36, 56 and 53 while COVID-19 mortality/ M is 3,3, 6 respectively.

Our study background hypothesis stands on possible heterogeneous immunity generated by malaria in addition to possible heterogeneous immunity generated by TB. This study will test COVID-19 mortality in malarious countries against TB prevalence standardized by BCG coverage and test the mortality again when malaria incidence effect is added to the composite sample and look for statistical associations and significances. This study addresses mortalities instead of morbidities as far as real number of affected people are beyond counting in view of many confounders affect testing and because of widely distributed asymptomatic persons. Furthermore, this study considers standardized TB prevalence to BCG coverage instead of non-standardized TB prevalence values.

## Material and methods

Data used are publically available as follows:

COVID-19 mortality taken on August,31 – September, 1^st^ 2020: https://www.worldometers.info/coronavirus/?

TB prevalence /100,000 : world bank site : https://data.worldbank.org/indicator/SH.TBS.INCD

Malaria incidence for 2018 /1000 :1- WHO. World Malaria Report 2019 at site : file:///C:/Users/zz/Downloads/9789241565721-eng.pdf

2- World bank site :https://data.worldbank.org/indicator/SH.MLR.INCD.P3

3- Max Roser and Hannah Ritchie (2013) - “Malaria”. *Published online at OurWorldInData*.*org*. Retrieved from: ‘https://ourworldindata.org/malaria’ [Online Resource]. Publically available at: https://ourworldindata.org/malaria

BCG data: WHO. BCG. Immunization coverage estimates by country publically available at: https://apps.who.int/gho/data/view.main.80500?lang=en

Data Availability: Datasets generated and/or analyzed during this study are shown in attached supplementary file.

### Patient and public involvement statement

It was not appropriate or possible to involve patients or the public in this work given that we used general practice level summated data and related publically published morbidity and mortality statistics and patients were not involved.

We did not include case fatality rate because no. of tests /Million vary from country to another country making COVID-19 cases/ 1 M is untrusted indicator. We took mortality /M as more trusted one.

All countries and territories under 1 million population were excluded to ovoid upgrade correction to 1 million by multiplication of figures these are : Sao Tome and Principe, Guyana, Djibouti, Suriname, Cabo Verde (Cape Verde), Burkina Faso, Belize, Bhutan, Comoros, Mayotte (France), Western Sahara, Vanuatu, Solomon Islands, Sao Tome and Principe.

Total number of malarious countries and territories enrolled in this study is 80. All of these countries have current BCG vaccination programs but differ vaccination coverage.

Countries with Zero malaria incidence are included as far as not yet get the malaria elimination certificate by WHO (Certification of malaria elimination is the official recognition by WHO of a country’s malaria-free status. WHO grants this certification when a country has proven, beyond reasonable doubt, that the chain of local transmission of all human malaria parasites has been interrupted nationwide for at least the past 3 consecutive years, and that a fully functional surveillance and response system that can prevent re-establishment of indigenous transmission is in place).^45^

### The Statistical Hypothesis

No significant effect due to standardized values of “TB/100000 by BCG Vaccination Coverage 2018 percent” on the COVID -19 Death /1 M. rates as a function factor with the presence of effectiveness an intermediate factor “Malaria incidence for 2018/1000” at the significance level greater than 0.05.

## Results and Findings

Hierarchical regression of composite -multiple linear model, was used for data analyses. The direct factor of reducing the mortality rates with reference to COVID-is the standardized TB/100,000 population by BCG vaccination coverage percentage in 2018 through dividing the factor of TB prevalence /100,000 population rates by the factor of BCG vaccination coverage in 2018, while the indirect effect that reduce the mortality rates, named as intermediate factor which is “The malaria incidence for 2018/1000 population”.

We investigated the validity of the assumptions of studied model that adopts the results of quantitative measurements. Table No. (1) shows the results of the multiple linear model fitness test resulted from the regression analysis of variance.

**Table (1):** Fitness tests resulted for the regression analysis of variance with and without effectiveness of an intermediate factor

| Model | SOV | SS | Df | MSS | F | P-value<sup>(*)</sup> |
| --- | --- | --- | --- | --- | --- | --- |
| Without Intermediate Factor | Regression | 170266.9 | 2 | 85133.45 | 3.833 | 0.026 S |
|  | Residuals | 1710329.0 | 77 | 22212.07 |  |  |
|  | Total | 1880595.9 | 79 | - |  |  |
| With Intermediate Factor | Regression | 322044.1 | 3 | 107348 | 5.235 | 0.002 HS |
|  | Residuals | 1558551.8 | 76 | 20507.26 |  |  |
|  | Total | 1880595.9 | 79 | - |  |  |

(\*) HS: Highly Sig. at P<0.01; S: Sig. at P<0.05.

The effectiveness of quality of model fitness is observed in the presence of the intermediate factor, the level of significance is greatly reduced compared to the case of the model quality in the absence of the intermediate factor.

On other hand, table No. (2) shows the results of estimating some descriptive statistics accompanying the analysis of the composite linear model.

The level of increase in the value of the multiple correlation coefficient is evidenced by the presence of the intermediate factor in the composite regression analysis.

In table (3) regarding the model of regression factor estimate (i.e. the standardized TB/100,000 by BCG vaccination coverage 2018), the table shows meaningful linear regression tested in two tailed alternative statistical hypothesis of studied factor playing effective role for reducing “COVID-19 death /1 M. population” as a function of the previous factor. Slope value estimation indicates that with increasing one unit of the studied factor, a negative influence on unit of the rates function factor occurred, and estimated as (−22.11). This decrement recorded significant influence at P<0.05. Other confounders are not included in model, i.e. “Constant term in regression equation” shows that non assignable factors which are not included in regression equation, ought to be informative, since estimation about (127) cases of “COVID-19 Mortality / 1 M.” expected initially without effectiveness of the restriction of studied factor.

Back to the results of the table No. (3), and regarding to composite regression estimate factors (i.e. the standardized TB/100000 by BCG vaccination coverage 2018 as direct effect, and malaria incidence for 2018/1000 population as indirect effect) results shows a meaningful composite linear regression tested in two tailed alternative statistical hypothesis of studied factor is playing effective role for reducing “COVID-19 deaths /1 M.” rates as a function of the previous factors. Slope value concerning that the first factor’s estimate indicates that with increasing one unit of the studied first factor, a negative influence on unit of the rates function factor would occur, and estimated as (−18.00). The decrement recorded significant influence at P<0.05, while with presence of indirect effect by the second factor, slope value indicates that with increasing one unit of that factor, a negative influence on unit of the rates function factor would occur, and estimated as (−0.331). The decrement recorded has highly significant influence at P<0.01. Other confounders that are not included in composite regression model, i.e. “Constant “ought to be informative, since estimated that about (152) cases of “Covid - 19 Mortality / 1 M.” expected initially without effectiveness of the restriction of studied factors.

## Discussion

We suggest that malaria is possible contributor to COVID-19 mortality in addition to TB.

Up to my knowledge previous studies regarding latent TB and BCG role in COVID-19 did not take malaria factor when testing prevalence of TB influence on COVID-19 morbidity or mortality. We think that could lead to bias. In this paper we test TB prevalence against COVID-19 mortality excluding non malarious countries. We think that this would exclude to certain extent the influence of non-malaria status on the sample tested. On other hand we test influence of TB in absent of malaria effect and in the presence of malaria effect as shown in statistical method. TB prevalence was adjusted to BCG coverage as far as *Mycobacterium tuberculosis* and *Mycobacterium Bovis* were suggested to have protective effect through inducing trained immune response. TB prevalence represents latent natural infection to certain extent since it represents 5-15 % of TB infection are presented as TB disease and 85- 95% as latent TB infection ^46^. The heterogeneous trained immunity is accumulative in nature since every year TB infects new targets^47^. BCG vaccination also exert accumulative effect since new persons (infants) are receiving the vaccine each year. Since the coverage rate represents the percentage of target population being vaccinated, we adjust TB prevalence figures to BCG vaccination coverage to present a single figure for statistical analyses representing both. We think testing TB influence by adjusting to BCG coverage in homogenous group of countries regarding malaria status will help us in avoiding selection bias if non malarious countries included. Selection bias is important thing to consider in research studies ^48^. In COVID-19 mortality studies we think that we have to consider many confounders including : BCG status, BCG coverage and malaria when we have to test effect of TB.

In correlation design, variables are measured simultaneously and cause effect relationship can’t look for so, but in some situations regression analysis can be used to infer causal relationships between the independent and dependent variables^49^ as far as theoretical and conceptual background link exists. In this study role of standardized TB prevalence and malaria being have been already explained before designing the model making background evidence for such testing.

Regression analysis is a set of statistical processes for estimating the relationships among variables i.e. ‘outcome variable’ and one or more ‘predictors^50,51^.

One type of regression analyses is Hierarchical multiple regression analysis that we choose for statistics for this study. It is defined as a subset of regression methods that we choose to proof theory of driven evidence for a proposed role of variables entered in blokes ^52^ (TB prevalence is 1^st^ in this study) and one additional variable in the 2^nd^ block ^52^ (malaria incidence in this study) on effect (reduction COVID-19 mortality here). This test allows us to look at the R^2^ change and F-statistic change between the two models, in addition to reporting the level of significance for each predictor variable which are entered into the model in pre-determined iterations^52, 53,54^. Table 2 shows that R –square was 0.091 increased to 0.171 when intermediate factor was added to composite.

**Table (2):** Results of some descriptive statistics accompanying the analysis of the composited linear model of a studied function.

| Model | R Multiple Corr. | R – Square | Adjusted R Square | Std. Error of the Estimate |
| --- | --- | --- | --- | --- |
| 1 | 0.301 | 0.091 | 0.067 | 149.0371 |
| 2 | 0.414 | 0.171 | 0.139 | 143.2036 |
| (1): Predictors: (Constant), BCG vaccination Coverage 2018 in percent, TB/100,000 |  |  |  |  |
| (2): Predictors: (Constant), BCG vaccination Coverage 2018 in percent, TB/100,000 , Malaria Incidence for 2018/1000 |  |  |  |  |

The first iteration will be with the most highly correlated variable to the outcome, that is standardized B prevalence here which is considered as dependent variable. Other variable that have some sort of association effect on the outcome to see what happens was malaria endemicity. Entry of this variable led to more significant increase in R-squared, then evidence of its reduction of COVID-19 mortality association was noted. It is found that TB prevalence gives a significant p value of 0.026 decreased to 0.002 when malaria endemicity added in 2^nd^ bloke. Parallel with decreased p value there is increase in correlation significant as evident by R and R square values. So we profe that Hierarchical multiple regression analysis gives us significant results to study the correlations of these variables with COVID-19 mortality.

The results show that the constant parameters (constants) constitute significant proportion causing COVID-19 mortality measured (152.38). TB prevalence when standardized to BCG coverage rate made -22.11 change and when malaria incidence was included in regression model (constants) made -0.331 further change. Constants constituted a considerable number not included in regression model (table 3). These unknown confounders might have attributed to other possible factors such as population density, genetic factors, environmental factors, social distances and population habits previous. We took BCG coverage, TB prevalence and malaria incidence just for one year. While effects of TB prevalence and malaria incidence are accumulative for years rather than one-year effect. Looking for these variables is strongly advised in future studies. In this study the prevalence of mycobacterium spp. (standardized to BCG vaccine coverae) exposure of the populations is negatively associated with COVID-19 deaths per million populations, this supports the previous mentioned studies^25,26, 27,28^

**Table No. (3):** Estimating parameters of the regression and composite regression models in the presence of the indirect effect of the intermediate factor

| Regression and Composite Regression Models | B | Std. Error | Stand. Beta | t-value | P-value<sup>(*)</sup> |
| --- | --- | --- | --- | --- | --- |
| (Constant) | 126.98 | 25.87 | - | 4.908 | 0.000 |
| Standardized of TB/100000 by BCG Vaccination Coverage 2018 percent | -22.11 | 8.53 | -0.282 | -2.593 | 0.011 |
| (Constant) | 152.38 | 26.58 | - | 5.734 | 0.000 |
| Standardized of TB/100000 by BCG Vaccination Coverage 2018 percent | -18.00 | 8.34 | -0.229 | -2.16 | 0.034 |
| Malaria Incidence for 2018/1000 | -0.331 | 0.122 | -0.289 | -2.717 | 0.008 |

(\*) HS: Highly Sig. at $P < 0.01$ ; S: Sig. at $P < 0.05$ .

Standardization TB prevalence standardization for BCG coverage is very important factor regarding studying countries currently implementing BCG programs as far as coverage is reflecting the degree of benefits added to the factor (latent TB prevalence) the coverage do and that’s what we do in this study. Likewise the influence of time duration of cessation of BCG vaccination program is a factor in determining COVID-19 mortality^55^ in countries ceased implementing this vaccine. BCG vaccination status for countries being of concern earlier in this pandemic before latent TB becomes of more concern but previous early studies were conflicting and were criticized because of possible confounding factors. This correlation although is statistical, but the theoretical bases for causal relation pave the way for further studies to proof TB and malaria roles and effects on reduction of COVID-19 mortality. Further studies are needed in this context. Our findings can explain the variances in COVID-19 mortality among different countries much deeper than TB and BCG whether as separated effects or integrated one effect.

Treatment of reservoir in TB and malaria control is slandered practice for elimination for these serious diseases.

Once concerns raised about their role in innate immune response is raised nowadays, does changing protocols is needed in next future if causation is proved on clinical grounds? Does the role of weighting the benefit risk ratio will be operated on leading to more conservative protocols in treating reservoirs ?

It is not too early to asking such questions but it is early to answer them unless further confirmatory studies done.

## Conclusions

Malaria, TB prevalence and BCG coverage rates are correlated factors in COVID-19 mortality.

## Supporting information

appendices/ malarous countries

## Data Availability

Data are publicly available.
links to publicly accessed  data are included within the manuscript

## Ethics and dissemination

Ethical permission is not necessary as this study analyzed publically published data and patients were not involved.

There is no conflict of interest.

## Acknowledgment

I am deeply grateful to Emeritus Professor Abdulkhaleq Abduljabbar Ali Ghalib Al-Naqeeb, Ph.D. in the Philosophy of Statistical Sciences at the Medical & Health Technology college, Baghdad-Iraq, for his assistance, support and supervision in data analysis and interpretations of finding results.

