## appendices/ malarous countries for "Influence of Malaria Edemicity and TB Prevalence / BCG Coverage on COVID-19 Mortality"

**Appendix 1: Reported Covid Deaths /M (31/8/2020-1/9/2020), by Country and Territory, TB prevalence /100,000, Malaria, BCG status, BCG Vaccination**

**Coverage 2018**

| **Region or country** | **COVID-19 Death /1 M** | **TB prevalence /100,000** | **Malaria**  **Incidence for 2018 /1000** | **BCG status**  **^Multiple BCG now= 3 or BCG NOW and booster just in the past =2. BCG only at birth setting =1^** | **BCG Vaccination**  **Coverage 2018** |
| --- | --- | --- | --- | --- | --- |
| [**Lao PDR**](https://data.worldbank.org/indicator/SH.MLR.INCD.P3?locations=LA) | **0** | **162** | **4.2** | **1** | **79** |
| [**Myanmar**](https://www.worldometers.info/coronavirus/country/myanmar/) **(Burma)** | **0.1** | **338** | **3.4** | **1** | **90** |
| [**Viet Nam**](https://www.worldometers.info/coronavirus/country/viet-nam/) | **0.3** | **182** | **0.1** | **1** | **95** |
| [**Niger**](https://www.worldometers.info/coronavirus/country/niger/) | **3** | **87** | **356.6** | **1** | **87** |
| [**Thailand**](https://www.worldometers.info/coronavirus/country/thailand/) | **0.8** | **153** | **0.4** | **2** | **99** |
| [**Uganda**](https://www.worldometers.info/coronavirus/country/uganda/) | **0.7** | **200** | **289.2** | **1** | **88** |
| [**Yemen**](https://www.worldometers.info/coronavirus/country/yemen/) | **19** | **48** | **45.8** | **1** | **64** |
| [**Angola**](https://www.worldometers.info/coronavirus/country/angola/) | **3** | **355** | **228.9** | **1** | **86** |
| [**Mozambique**](https://www.worldometers.info/coronavirus/country/mozambique/) | **0.7** | **551** | **305.4** | **1** | **95** |
| [**Mali**](https://www.worldometers.info/coronavirus/country/mali/) | **6** | **53** | **386.8** | **1** | **83** |
| [**Togo**](https://www.worldometers.info/coronavirus/country/togo/) | **3** | **36** | **267.3** | **1** | **83** |
| [**Benin**](https://www.worldometers.info/coronavirus/country/benin/) | **3** | **56** | **386.2** | **1** | **89** |
| [**South Sudan**](https://www.worldometers.info/coronavirus/country/south-sudan/) | **4** | **146** | **235.9** | **1** | **52** |
| [**Nigeria**](https://www.worldometers.info/coronavirus/country/nigeria/) | **5** | **219** | **291.9** | **3** | **53** |
| [**Rwanda**](https://www.worldometers.info/coronavirus/country/rwanda/) | **1** | **59** | **486.5** | **1** | **97** |
| [**Malawi**](https://www.worldometers.info/coronavirus/country/malawi/) | **9** | **181** | **213.6** | **1** | **92** |
| [**Malaysia**](https://www.worldometers.info/coronavirus/country/malaysia/) | **4** | **92** | **0.1** | **1** | **98** |
| [**Sudan**](https://www.worldometers.info/coronavirus/country/sudan/) | **19** | **71** | **46.8** | **1** | **88** |
| [**S. Korea**](https://www.worldometers.info/coronavirus/country/south-korea/) | **6** | **66** | **0.1** | **2** | **98** |
| **DPR Korea** | **0** | **513** | **0.4** | **1** | **96** |
| [**Ethiopia**](https://www.worldometers.info/coronavirus/country/ethiopia/) | **7** | **151** | **31.8** | **1** | **85** |
| [**Zimbabwe**](https://www.worldometers.info/coronavirus/country/zimbabwe/) | **14** | **210** | **51.0** | **1*** | **95** |
| [**Madagascar**](https://www.worldometers.info/coronavirus/country/madagascar/) | **7** | **233** | **82.4** | **1** | **70** |
| [**Indonesia**](https://www.worldometers.info/coronavirus/country/indonesia/) | **27** | **316** | **3.9** | **1** | **81** |
| [**Kenya**](https://www.worldometers.info/coronavirus/country/kenya/) | **11** | **292** | **70.1** | **1** | **95** |
| [**Zambia**](https://www.worldometers.info/coronavirus/country/zambia/) | **16** | **346** | **156.7** | **2** | **91** |
| [**Botswana**](https://www.worldometers.info/coronavirus/country/botswana/) | **3** | **275** | **0.6** | **1** | **98** |
| [**Guinea**](https://www.worldometers.info/coronavirus/country/guinea/) | **4** | **176** | **283.9** | **1** | **72** |
| [**Haiti**](https://www.worldometers.info/coronavirus/country/haiti/) | **18** | **176** | **1.6** | **1** | **83** |
| [**Cameroon**](https://www.worldometers.info/coronavirus/country/cameroon/) | **15** | **186** | **247.0** | **1** | **88** |
| [**Senegal**](https://www.worldometers.info/coronavirus/country/senegal/) | **17** | **118** | **55.8** | **1** | **83** |
| [**CAR**](https://www.worldometers.info/coronavirus/country/central-african-republic/) **(Central African Republic** | **13** | **540** | **347.3** | **1** | **74** |
| [**Afghanistan**](https://www.worldometers.info/coronavirus/country/afghanistan/) | **36** | **189** | **29.0** | **1*** | **78** |
| [**Guinea-Bissau**](https://www.worldometers.info/coronavirus/country/guinea-bissau/) | **17** | **361** | **123.3** | **1** | **91** |
| [**Gambia**](https://www.worldometers.info/coronavirus/country/gambia/) | **40** | **174** | **66.0** | **1** | **94** |
| [**Nepal**](https://www.worldometers.info/coronavirus/country/nepal/) | **8** | **151** | **0.4** | **1** | **96** |
| [**Pakistan**](https://www.worldometers.info/coronavirus/country/pakistan/) | **28** | **265** | **3.4** | **1** | **86** |
| [**Ghana**](https://www.worldometers.info/coronavirus/country/ghana/) | **9** | **148** | **224.3** | **1** | **98** |
| [**Venezuela**](https://www.worldometers.info/coronavirus/country/venezuela/) | **14** | **48** | **32.7** | **1** | **92** |
| [**Mauritania**](https://www.worldometers.info/coronavirus/country/mauritania/) | **34** | **93** | **39.4** | **1** | **90** |
| [**Bangladesh**](https://www.worldometers.info/coronavirus/country/bangladesh/) | **26** | **221** | **0.7** | **1** | **99** |
| [**Philippines**](https://www.worldometers.info/coronavirus/country/philippines/) | **33** | **554** | **0.2** | **3** | **75** |
| [**India**](https://www.worldometers.info/coronavirus/country/india/) | **48** | **199** | **5.3** | **1** | **92** |
| [**Namibia**](https://www.worldometers.info/coronavirus/country/namibia/) | **32** | **524** | **26.7** | **1** | **94** |
| [**Equatorial Guinea**](https://www.worldometers.info/coronavirus/country/equatorial-guinea/) | **59** | **201** | **269.0** | **1** | **63** |
| [**Papua New Guinea**](https://data.worldbank.org/indicator/SH.MLR.INCD.P3?locations=PG) | **0.6** | **432** | **184.5** | **1** | **69** |
| [**Gabon**](https://www.worldometers.info/coronavirus/country/gabon/) | **24** | **525** | **248.2** | **1** | **87** |
| [**Eswatini**](https://www.worldometers.info/coronavirus/country/swaziland/)  **(Swaziland)** | **78** | **329** | **0.8** | **1** | **98** |
| [**Guatemala**](https://www.worldometers.info/coronavirus/country/guatemala/) | **154** | **26** | **0.3** | **1** | **88** |
| [**Iran**](https://www.worldometers.info/coronavirus/country/iran/) | **257** | **14** | **0.1** | **2** | **99** |
| [**Mexico**](https://www.worldometers.info/coronavirus/country/mexico/) | **499** | **23** | **0.3** | **2** | **96** |
| [**Honduras**](https://www.worldometers.info/coronavirus/country/honduras/) | **189** | **37** | **0.1** | **1** | **94** |
| [**Ecuador**](https://www.worldometers.info/coronavirus/country/ecuador/) | **372** | **44** | **3.3** | **1** | **90** |
| [**Costa Rica**](https://www.worldometers.info/coronavirus/country/costa-rica/) | **85** | **10** | **000** | **1** | **92** |
| [**Argentina**](https://www.worldometers.info/coronavirus/country/argentina/) | **188** | **27** | **000** | **2** | **93** |
| [**Dominican Republic**](https://www.worldometers.info/coronavirus/country/dominican-republic/) | **157** | **45** | **0.1** | **1** | **99** |
| [**Bolivia**](https://www.worldometers.info/coronavirus/country/bolivia/) | **424** | **108** | **1.4** | **1** | **90** |
| [**South Africa**](https://www.worldometers.info/coronavirus/country/south-africa/) | **238** | **520** | **1.7** | **2** | **70** |
| [**Colombia**](https://www.worldometers.info/coronavirus/country/colombia/) | **386** | **33** | **8.5** | **1** | **89** |
| [**Brazil**](https://www.worldometers.info/coronavirus/country/brazil/) | **568** | **45** | **5.1** | **2** | **90** |
| [**Panama**](https://www.worldometers.info/coronavirus/country/panama/) | **463** | **52** | **0.2** | **1** | **99** |
| [**Peru**](https://www.worldometers.info/coronavirus/country/peru/) | **876** | **123** | **4.7** | **1** | **81** |
| [**Saudi Arabia**](https://www.worldometers.info/coronavirus/country/saudi-arabia/) | **108** | **10** | **000** | **1** | **98** |
| [**Tanzania**](https://www.worldometers.info/coronavirus/country/tanzania/) | **0.3** | **253** | **124.3** | **1** | **99** |
| [**Chad**](https://www.worldometers.info/coronavirus/country/chad/) | **5** | **142** | **164.8** | **1** | **59** |
| **DR Congo** | **3** | **321** | **319.8** | **1*** | **83** |
| [**Somalia**](https://www.worldometers.info/coronavirus/country/somalia/) | **6** | **262** | **34.3** | **1** | **37** |
| [**Sierra Leone**](https://www.worldometers.info/coronavirus/country/sierra-leone/) | **9** | **298** | **320.4** | **1** | **90** |
| [**Liberia**](https://www.worldometers.info/coronavirus/country/liberia/) | **16** | **308** | **361.5** | **1** | **92** |
| [**Nicaragua**](https://www.worldometers.info/coronavirus/country/nicaragua/) | **21** | **41** | **7.1** | **1** | **98** |
| [**Congo**](https://www.worldometers.info/coronavirus/country/congo/) | **14** | **375** | **235.1** | **1** | **81** |
| **Burundi** | **0.08** | **111** | **250.3** | **1** | **91** |
| **Cambodia** | **0** | **302** | **23.7** | **1** | **93** |
| **China** | **3** | **61** | **0** | **3** | **99** |
| **Ivory Coast (Côte d’Ivoire)** | **4** | **142** | **330.6** | **1** | **98** |
| **Algeria** | **34** | **69** | **000** | **1** | **99** |
| **El Salvador** | **112** | **0** | **70** | **1** | **81** |
| **Eritrea** | **0** | **89** | **28.9** | **1** | **97** |
| **Oman** | **134** | **6** | **0** | **1** | **99** |
| [**Timor-Leste**](https://www.worldometers.info/coronavirus/country/timor-leste/) | **0** | **498** | **0** | **1** | **95** |

*** possible history of booster is not ruled out because lack of information.**

**Appendix 2 references for Datta**

| **Category** | **References** |
| --- | --- |
| **Covid-19 deaths/M as it is in 31/8/2020- 1/9/2020** | **Worldmeter. Coronavirus**  **Accessed: 1/9/2020**  [**https://www.worldometers.info/coronavirus/?**](https://www.worldometers.info/coronavirus/?) |
| **Malaria Incidence / 1000 population** | **1-CDC Malaria Information and Prophylaxis, by Country [E]**  [**https://www.cdc.gov/malaria/travelers/country_table/e.html**](https://www.cdc.gov/malaria/travelers/country_table/e.html)  **2- World malaria report 2019. Geneva: World Health Organization; 2019. Licence: CC BY-NC-SA 3.0 IGO.**  **file:///C:/Users/zz/Downloads/9789241565721-eng.pdf**  **3- The World Bank.** [**Data**](https://data.worldbank.org/)**. Incidence of Malaria**  [**https://data.worldbank.org/indicator/SH.MLR.INCD.P3**](https://data.worldbank.org/indicator/SH.MLR.INCD.P3)  **4-WHO. Global Health Observatory data repository. By Category.** [**Malaria**](https://apps.who.int/gho/data/node.main.A1362?lang=en) **.**[**Cases**](https://apps.who.int/gho/data/node.main.A1363?lang=en)  [**https://apps.who.int/gho/data/node.main.MALARIAINCIDENCE?lang=en**](https://apps.who.int/gho/data/node.main.MALARIAINCIDENCE?lang=en)  **5-** [**Roser M , Ritchie**](https://ourworldindata.org/team) **H. Our World in Datta. Malaria . last revised in October 2019.**  [**https://ourworldindata.org/malaria**](https://ourworldindata.org/malaria)  **Accessed: 3/9/202** |
| **TB prevalence /100000** | [**https://data.worldbank.org/indicator/SH.TBS.INCD**](https://data.worldbank.org/indicator/SH.TBS.INCD) |
| **BCG data and information**  **A – General** | **1-WHO. Global Health Observatory data repository.** [**By category**](https://apps.who.int/gho/data/node.main) **>** [**Immunization**](https://apps.who.int/gho/data/node.main.A824?lang=en)**. BCG. Immunization coverage estimates by country.**  [**https://apps.who.int/gho/data/view.main.80500?lang=en**](https://apps.who.int/gho/data/view.main.80500?lang=en)  **Accessed 3/9/2020**  **2- Zwerling A, Behr MA, Verma A, Brewer TF, Menzies D, Pai M. The BCG World Atlas: a database of global BCG vaccination policies and practices. *PLoS Med*. 2011;8(3):e1001012. doi:10.1371/journal.pmed.1001012**  **3- WHO. Global Health Observatory data repository**  [**By category**](https://apps.who.int/gho/data/node.main) **>** [**Immunization**](https://apps.who.int/gho/data/node.main.A824?lang=en)**.BCG Immunization coverage estimates by country**  [**https://apps.who.int/gho/data/view.main.80500?lang=en**](https://apps.who.int/gho/data/view.main.80500?lang=en) |
